# Single-cell RNA-seq and V(D)J profiling of immune cells in COVID-19 patients

**DOI:** 10.1101/2020.05.24.20101238

**Authors:** Xiaoying Fan, Xiangyang Chi, Wenji Ma, Suijuan Zhong, Yunzhu Dong, Wei Zhou, Wenyu Ding, Hongyan Fan, Chonghai Yin, Zhentao Zuo, Yilong Yang, Mengyao Zhang, Qiang Ma, Jianwei Liu, Ting Fang, Qian Wu, Wei Chen, Xiaoqun Wang

## Abstract

Coronavirus disease 2019 (COVID-19) has caused over 220,000 deaths so far and is still an ongoing global health problem. However, the immunopathological changes of key types of immune cells during and after virus infection remain unclear. Here, we enriched CD3+ and CD19+ lymphocytes from peripheral blood mononuclear cells of COVID-19 patients (severe patients and recovered patients at early or late stages) and healthy people (SARS-CoV-2 negative) and revealed transcriptional profiles and changes in these lymphocytes by comprehensive single-cell transcriptome and V(D)J recombination analyses. We found that although the T lymphocytes were decreased in the blood of patients with virus infection, the remaining T cells still highly expressed inflammatory genes and persisted for a while after recovery in patients. We also observed the potential transition from effector CD8 T cells to central memory T cells in recovered patients at the late stage. Among B lymphocytes, we analyzed the expansion trajectory of a subtype of plasma cells in severe COVID-19 patients and traced the source as atypical memory B cells (AMBCs). Additional BCR and TCR analyses revealed a high level of clonal expansion in patients with severe COVID-19, especially of B lymphocytes, and the clonally expanded B cells highly expressed genes related to inflammatory responses and lymphocyte activation. V-J gene usage and clonal types of higher frequency in COVID-19 patients were also summarized. Taken together, our results provide crucial insights into the immune response against patients with severe COVID-19 and recovered patients and valuable information for the development of vaccines and therapeutic strategies.

## Main

Severe acute respiratory syndrome coronavirus 2 (SARS-CoV-2) has spread globally to cause the coronavirus disease 2019 (COVID-19) pandemic and over 240,000 deaths, and the number of cases of infection and death are still rising rapidly. Patients with COVID-19 typically exhibit symptoms of fever, dry cough, fatigue, difficulty breathing, headache, diarrhea, nausea, muscle and/or joint pain, pneumonia, etc^1-4^. Some severe COVID-19 cases include development of acute respiratory distress syndrome (ARDS) and damage to multiple organs ^1,4-6^. The rapidly developing single-cell sequencing technologies provide powerful tools for exploring immune cell heterogeneity as well as immunotherapy and drug discoveries^7-9^. Here, to investigate what roles lymphocytes play in defending against SARS-CoV-2 viral infections, we recruited 13 participants. In addition to 4 patients with severe symptoms, we also included 6 cured patients and 3 healthy people who were negative for the SARS-CoV-2 virus tests. Peripheral blood mononuclear cells (PBMCs) of each individual were isolated from whole blood, and magnetic separation was used to collect CD3-positive cells or CD19-positive cells to enrich the T lymphocyte or B lymphocyte populations, respectively, from PBMCs. Based on the timing of blood collection, the 6 recovered patients were divided into 2 groups: the blood samples of 3 patients were collected within one week after the diagnosis with negative results of the SARS-CoV-2 virus test and no clinical symptoms (which were categorized as recovered patients at the early stage (RE patients)), while the blood samples of the other 3 cured patients were collected 20 days after a negative diagnosis and hospital discharge (which were named recovered patients at the later stage (RL patients)). After CD3 or CD19 antibody selection using MCS separation, single-cell mRNA transcriptome and single-cell V(D)J sequences of T lymphocytes and B lymphocytes were collected and analyzed. In total, we obtained scRNA-seq (single-cell RNA-sequencing) data from 70,984 cells and V(D)J combination information from 24,307 T lymphocytes and 46,689 B lymphocytes. A total of 42,791 cells (15,134 T lymphocytes and 27,657 B lymphocytes) were identified with matched gene expression and V(D)J combination profiles at the single-cell level.

We first performed unbiased clustering of the single-cell mRNA profiles and identified 38 clusters, which could be categorized into 18 cell types (Fig. 1a-c, S1a). There were 9 classes of T lymphocytes (CD3E+) identified, including 4 subtypes of *CD4+* T cells (11,961 cells) and 5 subtypes of *CD8+* T cells (17,683 cells) (Fig. 1a, b). Interestingly, we observed that the percentage of T cells was decreased in SARS-CoV-2-infected individuals, especially in patients with severe symptoms, compared with uninfected individuals (Fig S1b). For the B lymphocytes, we identified 4 *CD19+* and *CD79A+* classes: B cells (22,728 cells), memory B cells (5,617 cells), plasma B cell (2,284 cells) and plasmablasts (698 cells) (Fig 1a-c). In addition to T and B lymphocytes, we also picked up some other types of PBMCs, including monocytes (7,508 *CD14+* monocytes and 777 *FCGR3A+* monocytes), dendritic cells (130 cells), natural killer (NK) cells (1,282 cells), and platelets (316 cells) (Fig. 1a-c, S2). These cell types showed a similar number of detected genes, except for platelets (Fig. S1c). The cells from the samples of the same group (healthy, RE, RL and severe groups) showed similar distributions, suggesting that patients at similar stages after virus infection might experience similar immune responses (Fig. S3a). Intriguingly, we observed enrichment of some cell types in some sample groups (Fig 1d, e, S3b). For example, the two types of T helper cells, which are reported to have different immune responses, showed distinct proportions in the four groups; specifically, Th2-like follicular helper (Tfh) cells (*ICOS*+), which specialize in activating B cells to produce antibodies for immune responses in defending the body from extracellular pathogens^10^, were largely enriched in recovered patients, especially in the RE samples (Fig 1e, S3b). While Th1 cells, which are responsible for cell-mediated immunity by activating macrophages^10^, showed a larger population in RL samples than in the other sample types, reduced populations of both types of T helper cells were detected in patients experiencing severe virus infection (Fig. S3b). Accordingly, the majority of B lymphocytes in RE samples were B cells (Fig S3b). The majority of plasma B cells (85.4%), which are responsible for antibody production in an effective immune response, in patients with severe clinical features (Fig 1d, e, S3b).

**Figure 1.**
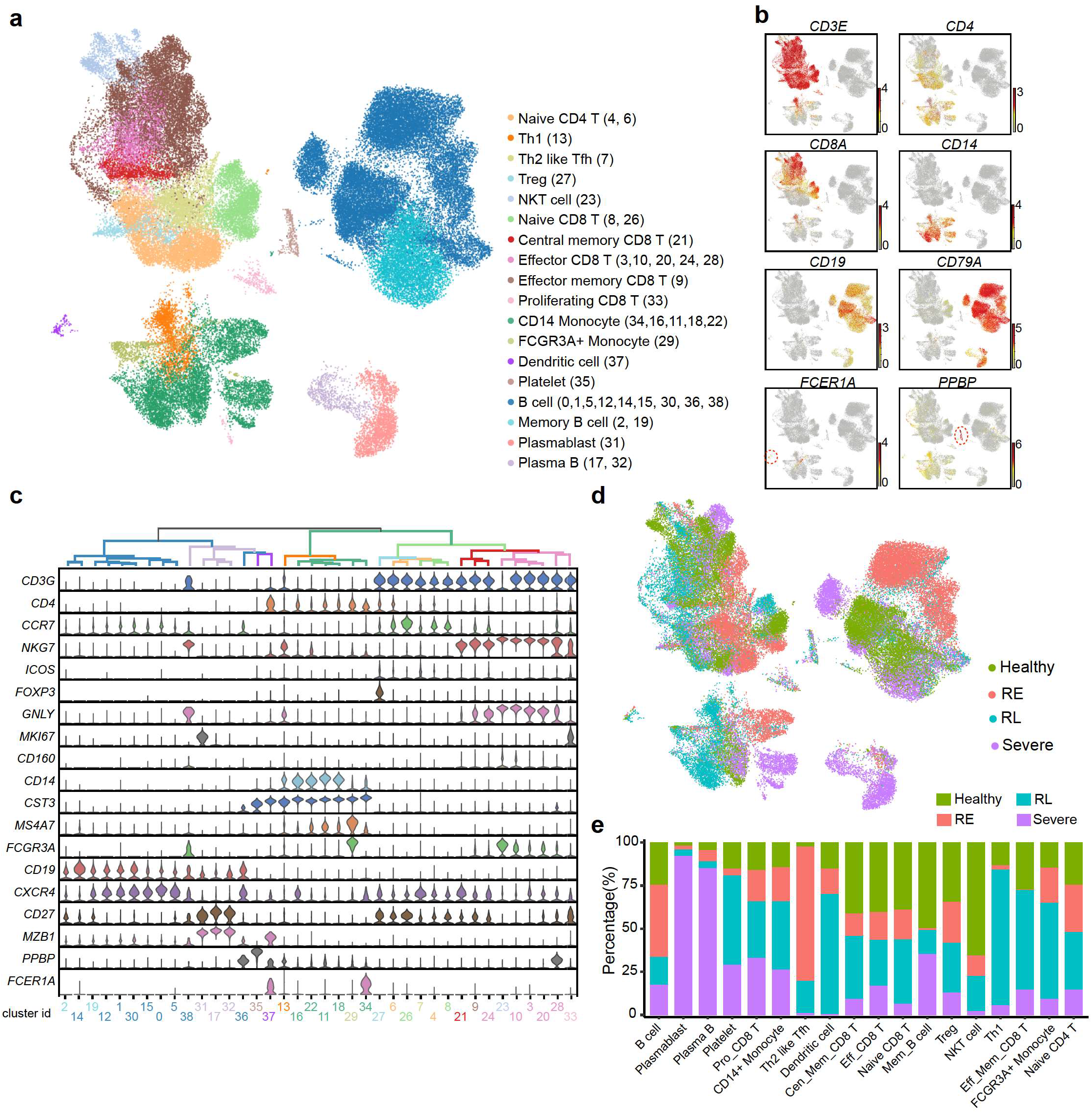
Cell type characterization of PBMCs in COVID-19 patients and normal controls. a. A total of 70,984 cells enriched with CD3 and CD19 antibodies were divided into 38 clusters, which were categorized into 18 cell types. The numbers in the brackets are the corresponding clusters for each cell type. b. Feature plot showing the identical markers for general cell types: *CD3E* is a general marker for T lymphocytes, which are further divided into *CD4+* and *CD8+* T lymphocytes. *CD19* and *CD79A* represent B cells; *CD14* is a marker for monocytes; *FCER1A+* represents dendritic cells; PPBP represents platelets. c. Histograms showing the relationships of the 38 clusters and marker genes for each cell type are shown with violin plots in each cluster. d. UMAP showing the cell type distribution of the four groups (healthy, RE, RL and severe) of patients. e. Bar plot showing the compositions of each cell type in healthy, RE, RL and severe patients.

To further investigate how T cells responded to SARS-CoV-2 virus infection, we first compared the T cell subclusters (Fig. S4a). We found that the 15 subclusters of T cells could be grouped into 5 modules based on the highly variable genes in the T cell transcriptomic profiles rather than on patient groups (Fig. S4a, b). Then, we analyzed the differentially expressed genes (DEGs) of T cells from different patient groups (Fig. 2a). We found that inflammatory genes, including *IFNG* (interferon gamma), *CD160, S100A8* and *GZMA*, were highly expressed by T cells from patients with severe infection, indicating that T cells were highly activated to participate in the response against virus infection in these patients (Fig. 2a-d). Accordingly, gene ontology (GO) analysis of the DEGs suggested that cytokine production and immune response-related leukocyte activation may occur in COVID-19 patients with severe symptoms (Fig. 2b). Interestingly, T cells from RE patients highly expressed *RNF125, CXCR4*, and *PELI1*, indicative of T cell activation still existing even after recovery at early time, which is consistent with the GO analysis results.

**Figure 2.**
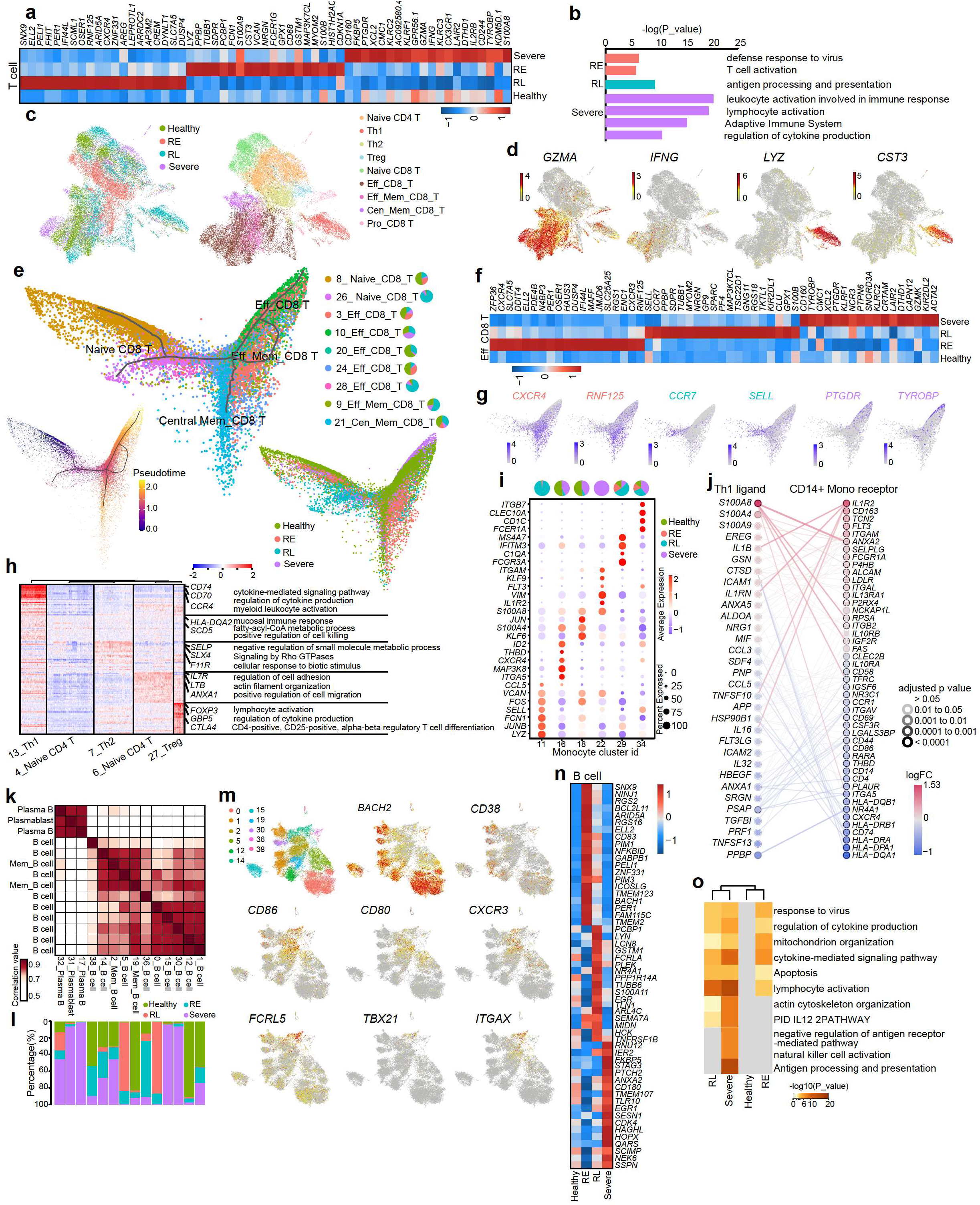
Novel cell subtypes and distinct gene regulation in COVID-19 patients. a. Heatmap showing the patient group DEGs in T lymphocytes. b. Identical GO terms enriched in the T lymphocytes of RE, RL and severe patients. c. UMAP showing the cell types of T lymphocytes and their patient group identities. d. Feature plot showing the patient group DEGs and cell type markers. e. Monocle map showing the differentiation of CD8 T cells upon SARS-CoV-2 infection. The patient group compositions for each cluster of CD8 T cells are shown by a pie chart. f. Heatmap showing the DEGs of effector CD8 T cells among healthy, RE, RL and severe patients. g. Feature plot of patient group-specific effector CD8 T cell genes on the monocle map, reflecting different stages of effector CD8 T cell differentiation in each group of patients. h. Heatmap showing the DEGs between the types of CD4 T cells. Representative genes and GO terms are shown on the right. i. Dot plots showing the expression of identical genes in subtypes of monocytes. The patient group compositions for each cluster are shown by the pie chart on the top. j. Ligand and receptor communication analysis of Th1 and *CD14+* monocytes between severe patients and other groups. k. Correlation of clusters of B lymphocytes based on gene expression patterns. l. Bar plot showing the composition of healthy, RE, RL and severe patients in the clusters of k. m. Feature plot showing the identical genes specifying B cell states and subtypes. n. Heatmap showing the DEGs of effector B cells among healthy, RE, RL and severe patients. o. Representative GO terms of group-specific genes in n.

Since CD8+ T cells, also known as cytotoxic T cells, play essential roles in recognizing, binding and killing cells when infected by viruses^11^, we determined the differentiation trajectories of the CD8+ T cells by monocle analysis^12^ (Fig. 2e, S5a). We found that the cells started from naïve CD8 T cells and then developed into central memory CD8 T, effector CD8 T and effector memory CD8 T cells, mirroring the classic CD8 T cell differentiation process activated by confrontation with pathogens. Intriguingly, we found two subgroups of naïve CD8 T cells (clusters 8 and 26) located in two different branches on the trajectory path, and their cell composition differed remarkably. The majority of cluster 8 cells were from the healthy group, while cluster 26 cells were predominantly from the RL group (Fig. 2e) and highly expressed *IL7R*, which is a receptor of IL7 and plays roles in early T cell development, homeostasis and activation. This indicated that naïve CD8 T cells from recovered patients may be at different states with specific transcriptome profiles (Fig S4c). Additionally, most central memory CD8 T cells were from healthy samples (41.4%), while most effector memory CD8 T cells (57.6%) were from RL samples, indicating that recovered patients probably have a recent memory of immune responses induced by SARS-CoV-2 infection. Next, we further analyzed the DEGs of effector CD8 T cells based on patient groups (Fig. 2f, g). The effector CD8 T cells from severe samples highly expressed *PTGDR*, which has been identified as a mediator of allergic airway inflammation^13^, and *GZMK*, *XCL2*, which were highly expressed in activated T cells, indicating that effector CD8 T cells were highly active in disease conditions. Additionally, *CXCR4* and *RNF125*, which play roles in T cell migration, maintenance and activation, were expressed by CD8 T cells from the RE group but not by those from the RL group (Fig. 2f, g), suggesting that inflammatory responses are still active when SARS-CoV-2 virus is eliminated. We also found that *CCR7* and *SELL* were relatively highly expressed by effector CD8 T cells from RL samples compared to those from other groups, indicating that these T cells may be at the point to transition into central memory T cells for long-term immune protection.

CD4+ T cells play critical roles in activating the cells of the innate immune system, B lymphocytes, and cytotoxic T cells for the immune response. We next analyzed DEGs of CD4+ T cell subtypes, including 2 subclusters of naïve CD4 T cells (clusters 4 and 6), and subclusters of Th1, Th2-like Tfh and Treg cells (Fig. 2h). The naïve CD4 T cells of cluster 6 and cluster 4 highly expressed genes related to cell adhesion/migration and positive regulation of cell killing, respectively, indicating that the two subgroups of naïve CD4 T cells may be in different states (Fig. 2h). Consistently, cluster 6 was enriched for naïve CD4 T cells from samples from recovered patients at late stages, and cluster 4 cells were dominantly from samples from recovered patients at early stages (Fig. S4a, b), indicating that some naïve CD4 T cells in the RE samples may have still been transitioning into T helper cells to participate in protection.

Although we used magnetic beads to enrich CD3+ and CD19+ lymphocytes, we still captured 8,285 monocyte, which were categorized as classical *CD14+* monocyte and non-classical *FCGR3A* (*CD16*)*+* monocyte (Fig 2i). The subclusters of monocytes showed distinct gene expression patterns that correlated with the status of SARS-CoV-2 infection (Fig. 2i, S5b). In *CD14+* monocyte, almost all cluster 22 cells (1,328/1,334) were from severe samples with high expression of *KLF6* and *IL1R2* (Fig 2i). As monocyte play a crucial role in the elimination of pathogens, which are activated by Th1 cells, we next analyzed the ligand-receptor reactions between Th1 cells and monocyte during defense against the virus. Cell-cell communication analysis of Th1 cells (ligands) and CD14+ monocytes (receptors) between the severe and other groups (healthy, RE, and RL groups) (Fig 2j) revealed that inflammatory signals *S100A8* and *IL1B, IL1RN*, and *IL16* on Th1 cells have stronger interaction with *ANXA2* and *IL1R2* on *CD14+* monocyte, respectively, while *CCL3/CCL5-CXCR4*--mediated signals and *TGFBI* signals were decreased in severe samples (Fig. 2j).

Together, these results show that although the numbers of T cells in the peripheral blood were reduced upon infection with the SARS-CoV-2 virus, the remaining T cells, especially *CD8*+ cytotoxic T cells, were activated, and their activities lasted for a while after recovery in patients, indicating that *CD8*+ T cells in the peripheral blood of COVID-19 patients play an important role in the immune response to the virus. We also observed a trend of transition from effector CD8 T cells to central memory T cells in recovered patients, which may be prepared for future protection from the SARS-CoV-2 virus.

B lymphocytes function in the humoral immunity component of the adaptive immune system by secreting the specific antibody to bind an antigen^14^. A total of 14 subtypes of B lymphocytes (including 9 clusters of B cells, 2 clusters of memory B cells, 2 clusters of plasma B cells and 1 cluster of plasmablasts) were grouped into 2 modules based on cell identities (Fig. 2k, l, S6a). We next examined the different subtypes of B cells (Fig. 2m). Clusters 0, 1, 12, 15, and 30 highly expressed the naïve B cell genes *BACH2* and *CD38*, while *CD80, CD86* and *CXCR3* were relatively highly expressed in clusters 2 and 38, indicating that these B cells may be activated. Notably, we found that the B cells of cluster 14 highly expressed *TBX21, FCRL5*, and *ITGAX*, marker genes of atypical memory B cells (AMBCs), which are induced by specific types of virus infection^15^.

In B cells, we found that the composition of each cluster differed in terms of infection status (Fig. 2l). To further explore the gene expression differences of B cells in the various COVID-19-related states, we next analyzed the DEGs and GO terms of virus-infected and healthy people (Fig2n, o, S6b). We found that genes playing roles in the response to viruses, the regulation of cytokine production, and apoptosis were enriched in patients with severe conditions (Fig. 2o). In addition, GO analysis indicated that the *IL12* signaling pathway was involved in regulating B cells in severe and recovered patients at the late stage (Fig. 2o). Genes related to natural killer cell activation and antigen processing and presentation were relatively enriched in B cells from samples taken from patients with severe symptoms, which is consistent with our observation that plasma B cells were high in that group (Fig 2o). Th2-like Tfh cells are considered to facilitate B cell activation by releasing inflammatory signals or via ligand-receptor interactions. Therefore, we further analyzed the ligand-receptor interactions between Th2-like Tfh cells and B cells among different groups (Fig S6c). Similar expression and interaction patterns of classical interleukins, such as *IL2, IL4, IL7*, and *IL10*, were observed among groups. However, cell-cell communications through *TNF* and chemokine signals differed among groups, indicating that Tfh cells may regulate B cells in a slightly different way at different stages of the immune response against SARS-CoV-2 (Fig S6c).

T cell receptor (TCR) and B cell receptor (BCR) characteristics are crucial for analyzing the T cell repertoires and B cell repertoires within samples from patients infected by SARS-CoV-2^16^. Thus, we explored the single-cell TCR and BCR V(D)J data in each group of samples. Interestingly, although the individuals with SARS-CoV-2 infection showed a reduced number of lymphocytes (Fig. S1b), we observed a high level of clonal expansion in both the TCR and BCR repertoires in severe samples, especially for the BCR repertoire (Fig. 3a, S7, S8). To understand the differences between expanding and nonexpanding B lymphocytes in response to virus, we further divided the B lymphocytes into monoclonal B lymphocytes and clonally expanded B lymphocytes. Transcriptome correlation analysis indicated that the monoclonal B lymphocytes in the four groups were similar, while the clonally expanded B lymphocytes were heterogeneous (Fig. 3b). We further analyzed the DEGs of the two types of B lymphocytes in patients with severe infection (Fig 3c). The clonally expanded B lymphocytes highly expressed *CD27, CD38, XBP1, MZB1, IFI6*, and *TNRSF17*, indicating activation and the effector functions of these B cells. To explore the preferential V and J combinations in COVID-19 patients, we first analyzed and listed the V and J combinations most frequently used in the BCRs and TCRs in all samples (Fig. 3d-e). Among these combinations, relatively frequent pairings of the BCR in RE patients were IGKV1-9::IGKJ4, IGHV2-70::IGHJ4 and IGHV3-33::IGHJ2, and the IGKV3-15::IGKJ1, IGHV3-53::IGHJ4, IGHV3-33::IGHJ1 and IGHV1-69::IGHJ4 combinations were frequent in RL patients. (Fig. 3d). In addition to these highly used pairs, IGLV2-23::IGLJ6 was a unique combination in severe COVID-19 patients (Fig 3d). Additionally, the TCR pairings with the highest frequencies in samples from early recovered patients were TRBV18::TRBJ1–1, TRBV4–1::TRBJ2–7, TRBV6–1::TRBJ1–5, TRAV14/DV4::TRAJ29, and TRBV20–1::TRBJ1–6, among others (Fig. 3e). We next compared the usage of BCR and TCR V(D)J genes in COVID-19 patients with that in healthy people (Fig. 3f, g). We identified a relatively high usage of IGHJ6, IGHV3-30, IGHV3-33, IGHV-40-2, IGKJ2, IGKV1D-39, and IGKV4-1 in COVID-19 patients compared to healthy people (Fig. 3f). We then analyzed and revealed the amino acid sequences of the CDR3 gene in high-frequency TCR clones in different samples (Fig. 3h). These recognition sequences may have special functions during SARS-CoV-2 infection.

**Figure 3.**
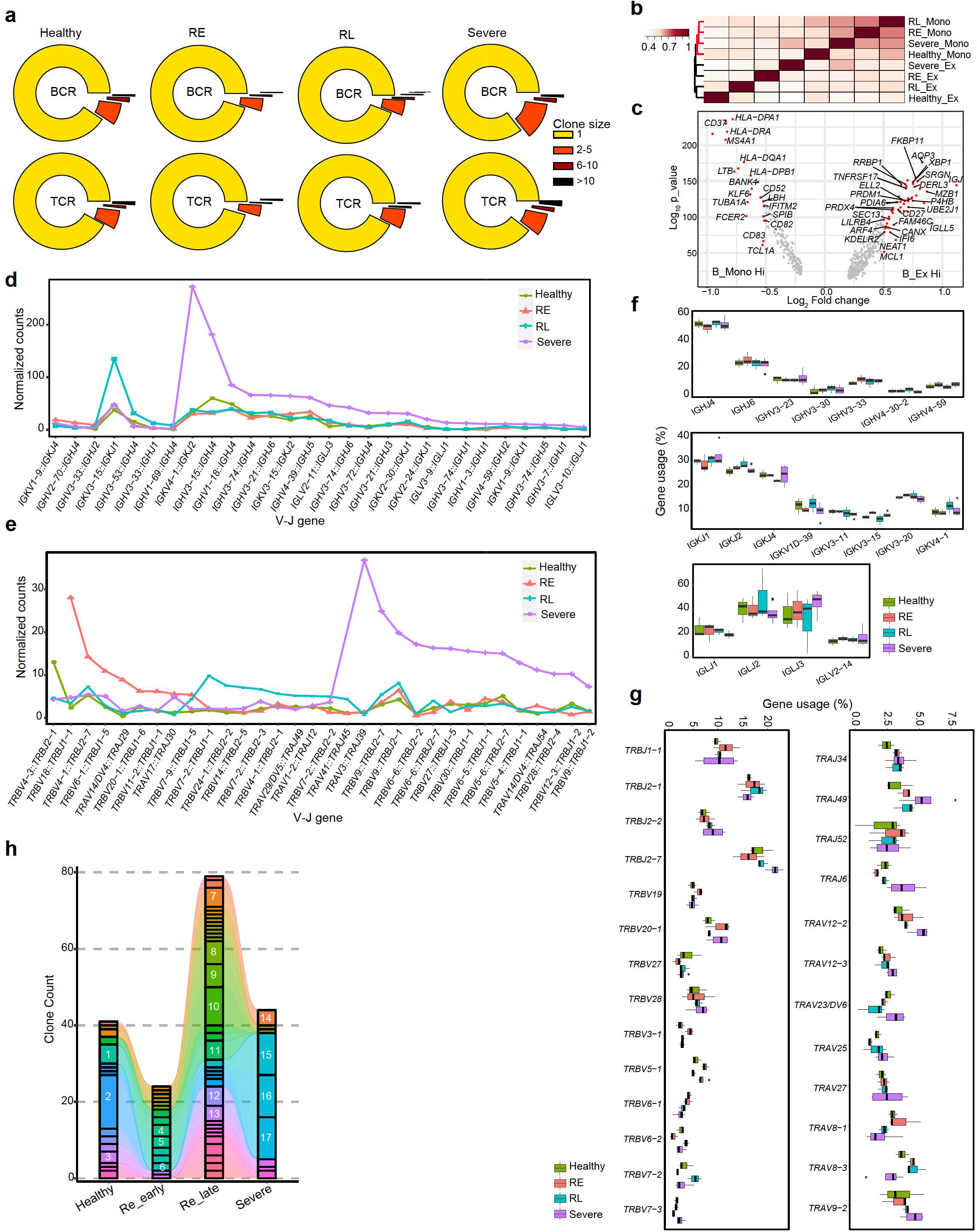
BCR and TCR clone expansion in COVID-19 patients. a. Calculation of expanded clone ratios of BCRs and TCRs in healthy, RE, RL and severe patients. b. Gene expression correlation analysis among monoclonal B cells and clonally expended B cells in the four patient groups. c. Volcano plot showing the DEGs of monoclonal B cells and clonally expended B cells in the severe patients. The latter showed a high activation state, expressing genes such as *XBP1, MAZB, IGLL5*, etc. d, e. The frequent VJ gene combinations in BCRs and TCRs within each group of COVID-19 patients. f,g. The V and J genes showing high usage frequency in BCRs and TCRs of COVID-19 patients. h. TCR clone type tracking showing the enriched CDR3 types (only those types with a clone size >1 are shown here) in healthy, RE, RL and severe patients.

Since we had both scRNA-seq transcriptome and TCR/BCR data, we next integrated and analyzed 42,791 cells (15,134 T lymphocytes and 27,657 B lymphocytes) with both types of information. All lymphocytes showed different degrees of clone expansion, and the effector CD8 T cells of severe COVID-19 patients showed relatively high clone numbers (Fig. 4a, b, S9). The plasmablast and plasma B cells in severe COVID-19 patients showed the highest degree of clonal expansion, indicating that SARS-CoV-2 infection may induce B cells to differentiate into plasma B cells to secrete antibodies against the virus. Hence, we analyzed the developmental trajectory of plasmablast and plasma B to trace the cell lineage. Interestingly, we found that plasmablast differentiated in two directions (cluster 17 and cluster 32) with distinct gene expression patterns (Fig. 4c, d). A total of 98.8% of the plasma B cells of cluster 17 were from severe COVID-19 patients, while cluster 32 consisted of plasma B cells from all groups (Fig. 2m). The tracing of BCR clones revealed a preferential differentiation of plasmablast into plasma B cells during SARS-CoV-2 infection (Fig. 4c). Further comparison of the two clusters of plasma B cells revealed that cells of cluster 17 highly expressed *FOS, IFI6, IGLL5* and*MX1*, indicating that these cells were activated (Fig. 4d). Importantly, we traced the source of these active plasma B cells as AMBCs (cluster 14 B cells) with a specific clonotype, indicating the important role of AMBCs in the immune response to SARS-CoV-2. Together, these findings show that the BCR clonotypes enriched in the plasma B cells might be helpful for vaccine and antibody production.

**Figure 4.**
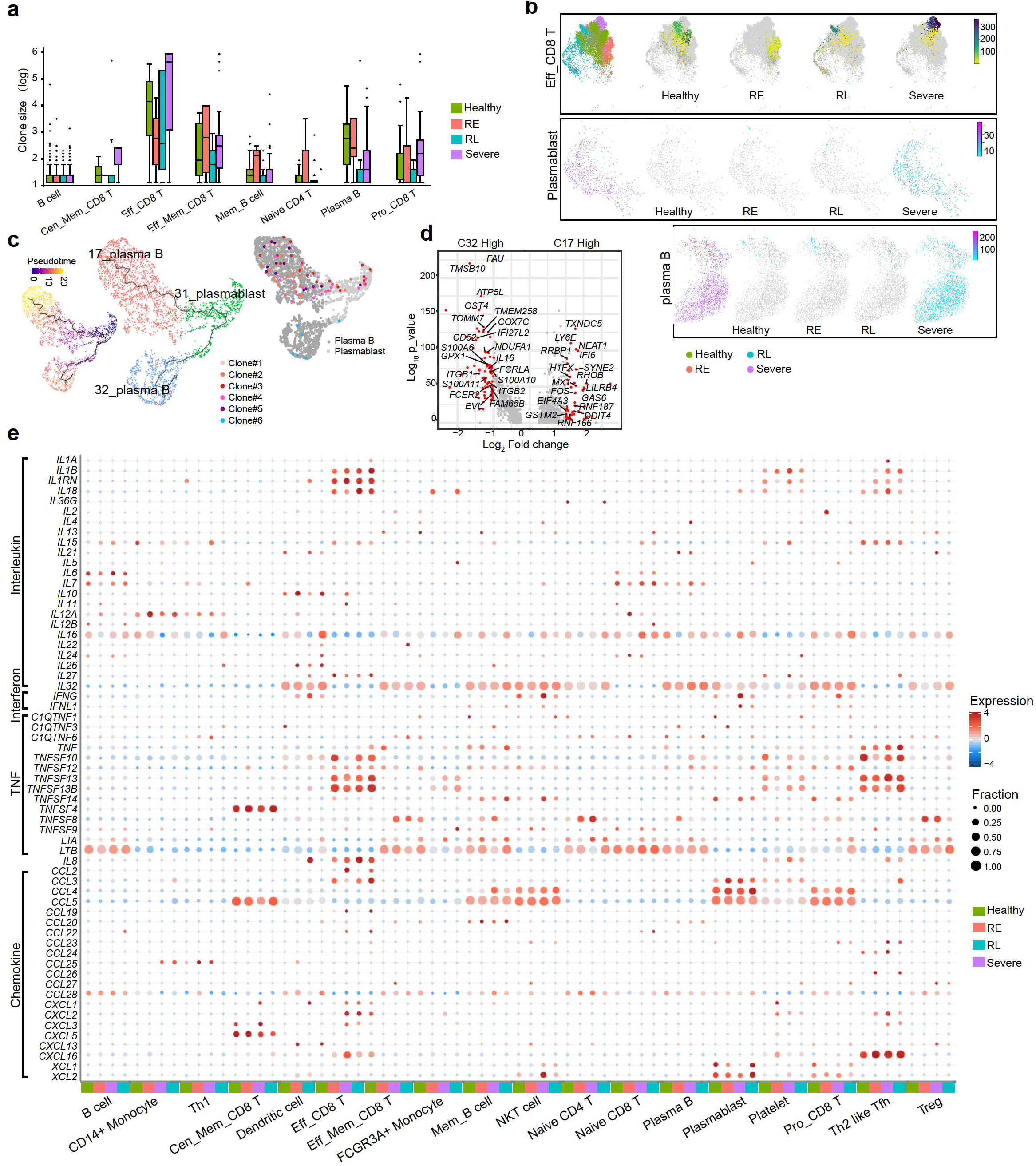
Cell type-based BCR and TCR clone expansion in COVID-19 patients. a. The clone size distribution of B cell and other cell subtypes in healthy, RE, RL and severe patients. b. Feature plot showing the clonally expanded cells and their clone size in effector CD8 T cells, plasmablasts and plasma B cells in healthy, RE, RL and severe patients. The effector CD8 T cells in severe samples showed extreme levels of clonal expansion. The plasmablasts had the highest fraction of cells showing clonal expansion. The plasma B cells in all groups were detected to be clonally expanded. The dark gray dots represent the cells with only one clone detected. c. Monocle analysis of plasmablasts and plasma B cells. Several clonotypes supporting the trajectory are shown. These clonotypes were all detected in severe patients. d. Volcano plot showing the DEGs of the two clusters of plasma B cells. e. Bubble plot showing the major chemokine genes expressed in each cell type of healthy, RE, RL and severe patients.

COVID-19 patients sometimes experience a cytokine storm, which forces the patient’s immune system into overdrive and can lead to death^17,18^. Therefore, we next focused on the global expression of cytokines in different cell types under various COVID-19-related statuses (Fig. 4e). *IL1A* and *IL1B* expression was high in *FCGR3A+* monocytes from severe COVID-19 patients. *IL6*, an inhibitor of which has been shown to ameliorate severe symptoms caused by cytokine release in patients with SARS-CoV-2 infections, was highly expressed by B cells, especially those from severe samples. Interestingly, recovered patients also have high expression of some interleukin molecules. We found that *IL12A* was high in B cells, memory B cells and plasma B cells from RE samples. *IL16* was highly expressed by effector memory CD8 T cells, naïve CD4 T cells, proliferating CD8 T cells and plasma B cells from RL samples. In addition to interleukins, *IFNG*, which is crucial for immunity against intracellular pathogens, was highly expressed by many types of CD8 T cells (effector CD8 T, effector memory CD8 T, naïve CD8 T and proliferating CD8 T cells) from severe patients. In addition, *TNF* (tumor necrosis factor), as an activator of the immune system, was highly expressed by monocytes and central memory CD8 T cells from RL samples. In terms of the expression pattern of chemokines, *IL8/CXCL8* expression was high in monocytes, Th1 cells and proliferating CD8 T cells from severe samples. *CXCL2* was generally expressed by classical and non-classical monocytes as well as Th1 cells from infected and recovered samples. Overall, the differential expression of interleukins, interferons, growth factors and chemokines in different types of cells in different SARS-CoV-2 infection conditions suggests that the immune system might work in slightly different ways in patients during infection and recovery at early or late stages.

In summary, we have illustrated changes in lymphocyte characteristics, including cell type, inflammatory status, gene expression and V(D)J recombination sequence, upon SARS-CoV-2 infection and after recovery. Coordinated and effective responses by innate and adaptive immune cells are crucial for body protection and virus clearance. Our results have revealed an active inflammatory response not only in severe COVID-19 patients but also in recovered patients at an early stage (within one week after diagnosis with negative results of the SARS-CoV-2 virus test). Lymphopenia is a common feature in severe COVID-19 patients, including decreased CD4+ T cells, CD8+ T cells, B cells and natural killer (NK) cells ^4,19,20^. In our study, we found that the proportion of T cells was reduced whereas the monocyte ratio was increased in severe COVID-19 patients compared to healthy people. However, CD8+ T cells, which are crucial for directly attacking and killing virus-infected cells, were active and highly expressed inflammatory genes, such as *GZMA* and *INF*, in severe COVID-19 patients as well as RE patients. Studies have revealed that patients who recovered from SARS developed specific memory T cells, which were still detectable up to 2 years after recovery^21,22^. Our results indicate that a group of effector CD8+ T cells may be at the stage of the process of transforming into central memory T cells in recovered patients at late stage (20 days after diagnosis with negative results of the SARS-CoV-2 virus test). It is very possible that these memory T cells could be vital for protecting these people from SARS-CoV-2 virus reinfection.

B cell responses accompanied by CD4+ T follicular helper cell responses were observed in COVID-19 patients. In B cells, genes related to the response to viruses, the regulation of cytokine production, and apoptosis were enriched the most in patients with severe infection and to a slightly lower degree in recovered patients. Consistent with the high ratio of plasma cells in severe patients, genes related to antigen processing and presentation were also highly expressed in B lymphocytes of these patients. The plasma cells of COVID-19 patients exhibited highly expanded clones. Notably, when tracing the BCR clones, we found that the source of a subtype of plasma cells in severe COVID-19 patients was atypical memory B cells (AMBCs), which are a unique subcluster of B lymphocytes, indicating that the B cells were highly active in the immune response against SARS-CoV-2. Recent studies have suggested that antibody-dependent enhancement (ADE) might be induced by SARS-CoV-2 infection in some cases^23^. Our results cannot determine whether ADE occurred in the patients with severe infection involved in this study. In the clinic, convalescent plasma has been used for the treatment of COVID-19, which potentially offers specific anti-SARS-CoV-2 polyclonal antibodies and has some positive impacts on patients^24,25^. Recent studies have reported that it is very likely that a subset of patients may not develop long-lasting antibodies to SARS-CoV-2. In our study, the numbers of plasma cells from recovered patients were quite limited. Whether these recovered patients are able to produce an immune response against future SARS-CoV-2 encounters needs further investigation.

One function of the adaptive immune system is recognizing and remembering specific pathogens through T cell responses and antibodies produced by plasma cells^26,27^. Since COVID-19 is a pandemic, many efforts are underway to develop therapeutic approaches against SARS-CoV-2 worldwide. One strategy is inducing SARS-CoV-2-specific memory CD8 T cells from a vaccination. These memory CD8 T cells can differentiate into effector T cells to kill infected cells before they produce mature virions when the body is truly attacked by SARS-CoV-2. Another strategy is to develop therapeutic antibodies against SARS-CoV-2 using BCR sequences from recovered patients^16,27-31^. We analyzed the characteristics of TCRs and BCRs from patients with severe infection and patients who had recovered and revealed the cell-type specific V(D)J sequences enriched in each group. This information provides valuable resources for the development of vaccines and antibodies for COVID-19 immunotherapies.

## Data Availability

The scRNA-seq data and V(D)J sequencing data used in this study have been deposited in the GSA.

## Acknowledgements

This work was supported by the special project for COVID-19 of Guangzhou Regenerative Medicine and Health Guangdong Laboratory (2020GZR110106009), National Basic Research Program of China (2019YFA0110100).

## Author Contributions

W. C, Q.W., X.W. and X. F. conceived the project, designed the experiments. X. C., Y, D., L. Y. and M. Z. performed the sample preparation and single-cell RNA sequencing experiment. H. F., T. F. and C. Y. helped on data transfer. X. F, W. M. and S. Z analyzed the mRNA data. W. Z., W. D, Q. M. and J. L. analyzed the V(D)J data. W. Z., S. Z. and Z. Z. performed clone tracing analysis. Q. W. and X. F wrote the manuscript. All authors edited and proofed the manuscript.

## Methods

### Sample collection and single cell library preparation

This study was approved by the Medical Ethics Committee of Wuhan Infectious diseases hospital, China and the blood samples were collected from patients who had signed the informed consent. The Human Peripheral Blood Mononuclear Cells (PBMCs) were obtained by Ficoll density gradient centrifugation (DakeweBiotech) according to the manufacturer’s instruction and then applied to the red blood cell lysis solution (Miltenyi Biotec) for 10 minutes at room temperature. Following washing steps with PBS containing 2% FBS, T cells were isolated from PBMCs using human CD3 microbeads (Miltenyi Biotec) and B cells were isolated using human CD19 microbeads (Miltenyi Biotec). Briefly, PBMCs were filtered using 30 αm nylon mesh (BD bioscience), mixed with CD3 and CD19 MicroBeads for 15 minutes at 4 °C and applied onto MACS column, respectively. After washing the column on MACS rack with MACS buffer, the columns were removed from the rack and the targeted T cells were eluted from the columns using PBS. Then the sorted B cells and T cells were immediately used for single cell RNA and V(D)J library preparation using Chromium Single Cell V(D)J Reagent Kits (10X Genomics) according to the manuals.

### ScRNA-seq data preprocessing

The raw sequencing data were aligned, quantified, called, and aggregated using the Cell Ranger Single-Cell Software Suite (version 3.1.0, 10x Genomics) count against the GRCh38 human reference genome with default parameters. The gene-cell counts matrix for the different batches of samples were aggregated by cell ranger by aggr with sequencing batch correction function on. Cells that passed the following filtration were kept for downstream analysis: gene number between 200 and 5000; UMI counts above 1000; percentage of UMIs from mitochondrial genes below 10% and that from hemoglobin genes below 1%, respectively.

### Dimension reduction and clustering

Scanpy^32^(V1.4.4) package was used to perform preprocessing of the scRNA-Seq data. The filtered gene-cell matrix was normalized to 10^4^ molecules per cell for sequencing depth with normalize_total function and log transformed with log1p function. The data variation caused by number of obtained UMI counts and percentage of UMIs from Mitochondrial was regressed out with the regress_out function. We obtained 1520 variable genes with mean expression ranging from 0.0125 and 3 and dispersion greater than 0.5. Uniform manifold approximation and projection (UMAP)^33^ was performed with the first 50 principal components from principal component analysis (PCA) for visualization of the single cells. Cell clustering was performed by Leiden algorithm^34^ (faster than Louvain algorithm and uncovers better cell partition) with resolution 1.8. 39 clusters were identified in total, among which cluster 0 and 25 were highly resembling each other according to our following cluster analysis. Therefore, we combined cluster 0 and 25 into one cluster labeled as cluster 0 throughout the analysis in this work.

### Differentially expressed genes (DEGs) analysis for single cell group

DEGs analysis between cell types/subject group/cell clusters was performed by Wilcoxon rank-sum test with FindAllMarkers function in Seurat^35^ (V3.0) with default parameters. Genes with expression natural log fold change > 0.5 and Bonferroni correction adjusted p value < 0.01 were reported as significant DEGs. The top 300 genes ordered by absolute value of natural log fold change will be regarded as marker genes for a cell group if there are too many reported significant DEGs. Enrichment analysis of the marker genes were performed with Metascape^36^ (https://metascape.org).

### Single cell development trajectory reconstruction

The Monocle 3 package(V3.2.0) were applied to construct single cell pseudo-time trajectory to discover differentiating transitions^12,37,38^. We used highly variable genes identified by Seurat to sort cells in pseudo-time order. The actual precursor determined the beginning of pseudo-time in the first round of “orderCells”. UMAP was applied to reduce dimensional space and the minimum spanning tree on cells was plotted by the visualization functions “plot_cells” for Monocle 3.

### Cell-cell communication

Cell–cell communication was predicted by a method similar to that described by Kirouac et al^39,40^. We created a cell-cell communication interactome with known protein-protein interactions between receptor and ligand collected by Rubin et al.^40^. The involved gene list was further manually filtered with the DEGs of different states in our cell types. To investigate state-related perturbations in these putative cell–cell interaction networks, DEGs metrics (e.g., fold change, p value) from the MAST analysis in Seurat were used to build subnetworks for each set of interactions between cell types. In these networks, nodes represent ligands or receptors expressed in the denoted cell type, and edges represent protein–protein interactions between them. Nodes were colored to represent the magnitude of DGE. These values were scaled per cell type and summed to determine edge weight.

We used CellPhoneDB V2.0^41^ to calculate ligand-receptor interactions between T cells and B cells in different states. We used the sequencing depth normalized raw UMI counts as input into “cellphonedb method statistical_analysis” to analyze the dataset and select the significant pairs of ligand-receptor to plot.

### Evaluation of cytokine expression level in specific cell type across subject groups

We curated a list of genes that can produce cytokine, including chemokine, TNF, interferon and interleukin, which can potentially reflect the level of cytokine storm in the patients. For each cell type in each subject group, we applied a similar strategy to aggregate expression level and expression fraction in a cell population proposed by Peng et al., 2019^42^ and calculated two matrices ***E*** and ***F***. For each gene *g* and a cell type in a subject group *cg*, ***E****(g, cg)* is the mean expression of cells with positive expression and ***F****(g, cg)* is the fraction of cells in a cell group *cg* that have positive number of UMIs of g. The evaluation expression score for each gene in each cell type of each subject group is calculated as the product of the two matrices: ***Score = E^*^F***. Then we visualized this score with bubble matrix plot.

### TCR/BCR V(D)J sequencing and analysis

TCR/BCR V(D)J segments were enriched from amplified cDNA from 5’ libraries via PCR amplification using a Chromium Single-Cell V(D)J Enrichment kit according to the manufacturer’s protocol (10x Genomics). The FATAQ files for each single T cell were assembled by Cell Ranger vdj pipeline (v3.1.0), calling to the identification of CDR3 sequence and the rearranged V(D)J gene.

In order to get the dominant TCR/BCR of a single cell, we filtered a total of 70,996 high-confidence contig sequences in cell barcodes as follows: 1) kept the barcodes which was productive and marked as raw clonotype. 2) only TCR alpha-beta (TRA-TRB, we dropped slight number of TRA-TRG) or BCR heavy-light (IGH-IGL, IGH-IGK) paired chain were considered. 3) if more than one TCR/BCR paired chains were identified in one cell, we only kept the dominant paired chain (supported by largest number of UMIs) for it. Finally, we got a total of 57,932 cells with paired chain information (16,745 T lymphocytes and 41,187 B lymphocytes). A unique clonotype was defined by consistent CDR3 amino acid sequence, V gene and J gene. Chao1 repertoire diversity ^43^ and repertoire overlap analysis was estimated by VDJtools^44^. We used immunarch(V0.5.5)^45^ to compute gene usage against IMGT database (http://www.imgt.org/IMGTrepertoire/LocusGenes/). TCR clonotypes was annotated with VDJdb ^46^ antigen categories database.

### Data availability

The scRNA-seq data and V(D)J sequencing data used in this study have been deposited in the GSA (Genome Sequence Archive in BIG Data Center, Beijing Institute of Genomics, Chinese Academy of Sciences).

---

**Figure S1.**
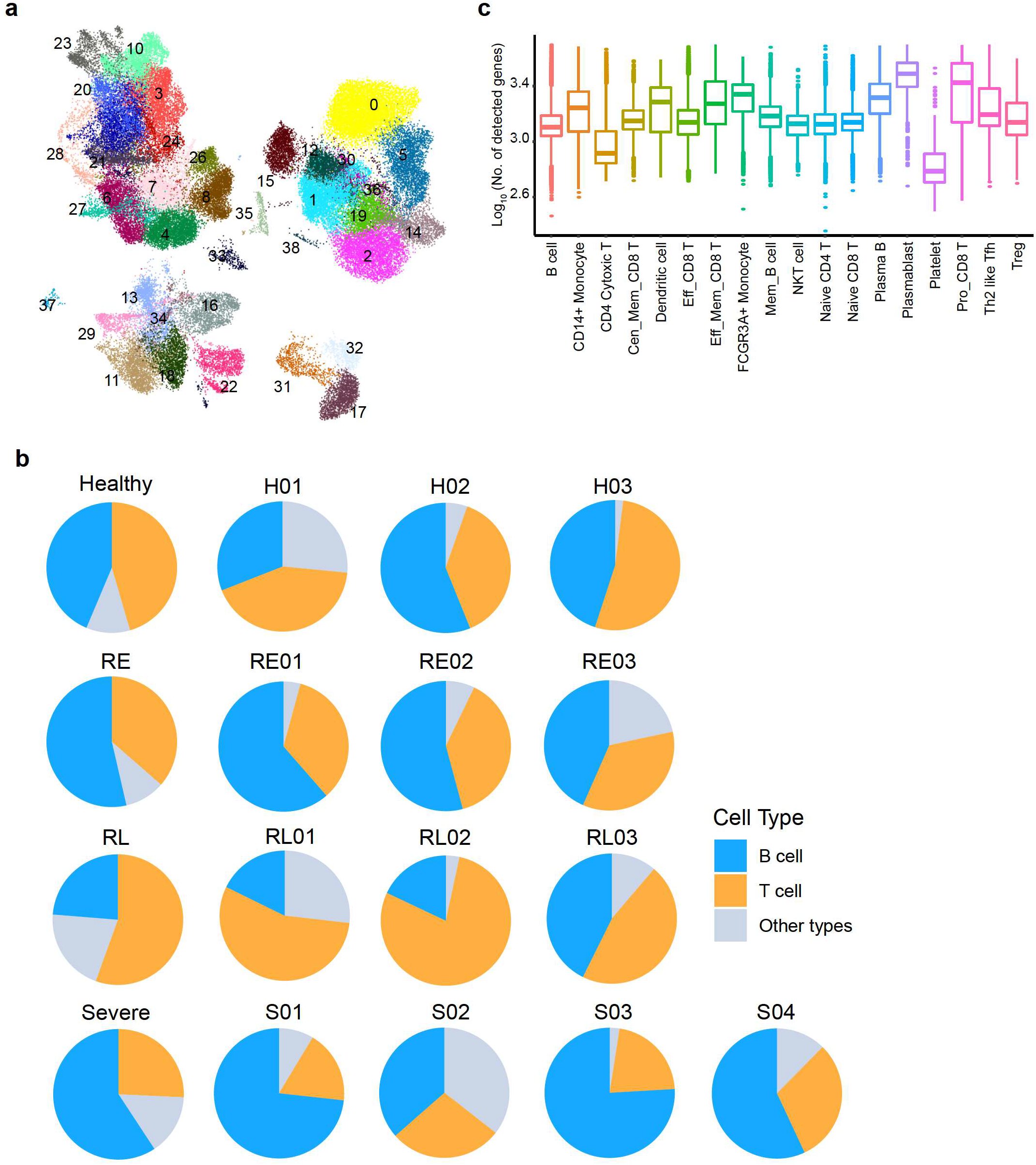
Clustering of single cells using scRNA-seq data. a. The 38 clusters of the 70,984 cells from 13 individuals. b. Pie chart showing the composition of cell in each group of samples. c. Barplot showing the gene number detected in each cell type.

**Figure S2.**
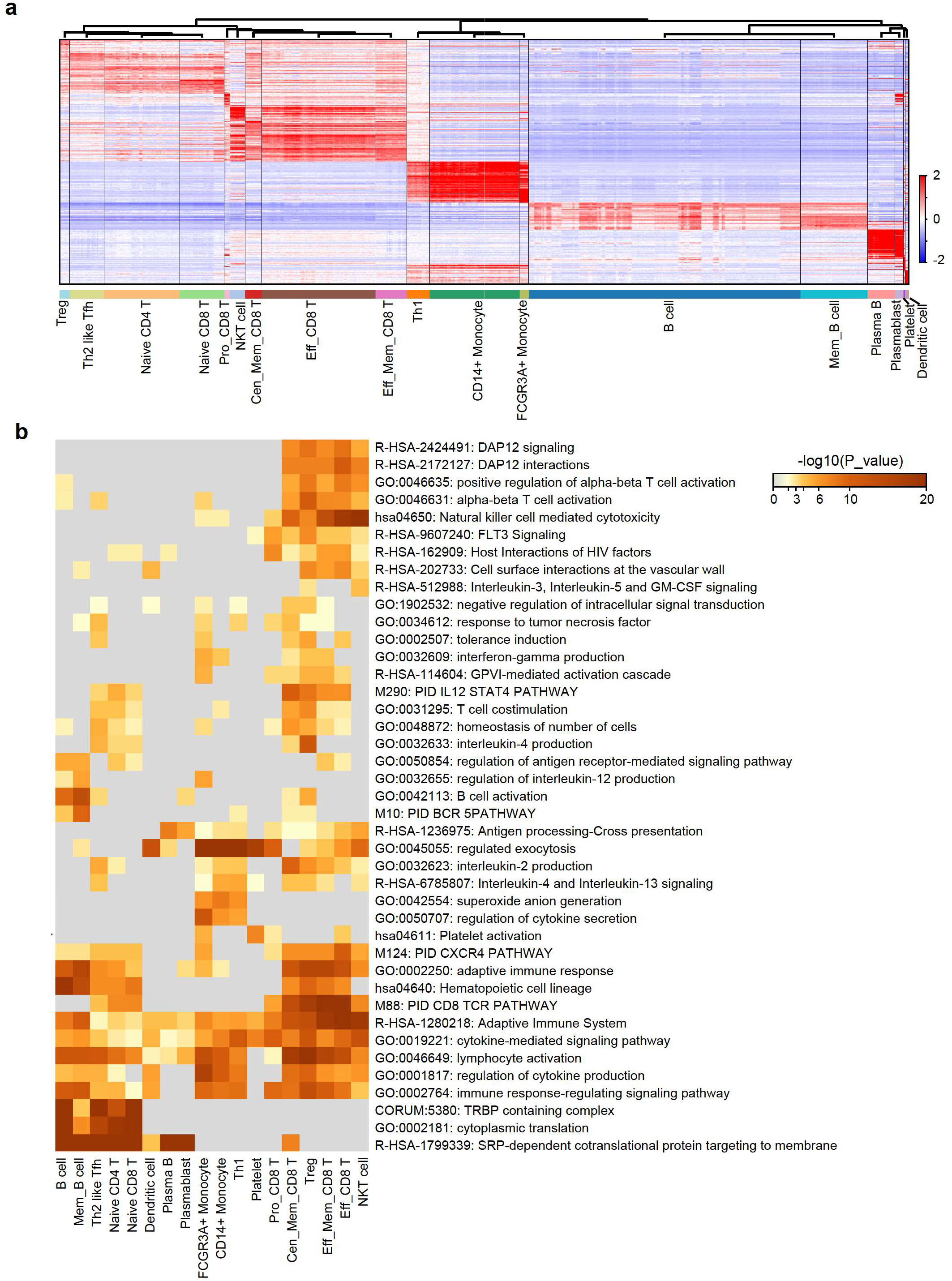
Cell types of CD3 and CD19 enriched PBMCs in COVID-19 patients. a. Heatmap showing the 18 cell types with specific gene expression patterns. b. The enriched GO terms of the cell type DEGs for the 18 cell types.

**Figure S3.**
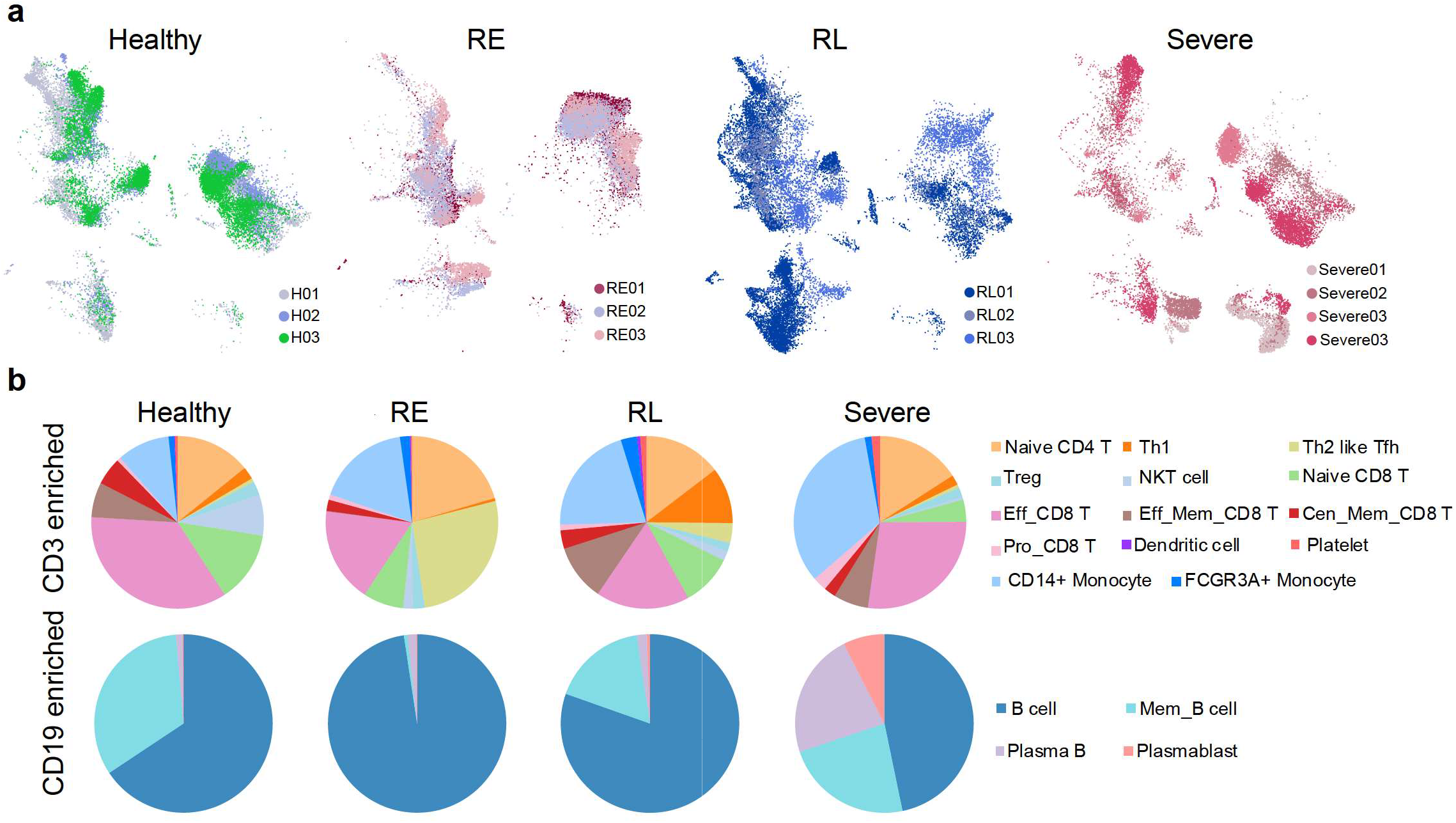
Cell types in each group of patients. a. UMAP showing the well replication of cells from different individual patients of the same group. b. Pie chart showing the cell type ratios in each group of patients.

**Figure S4.**
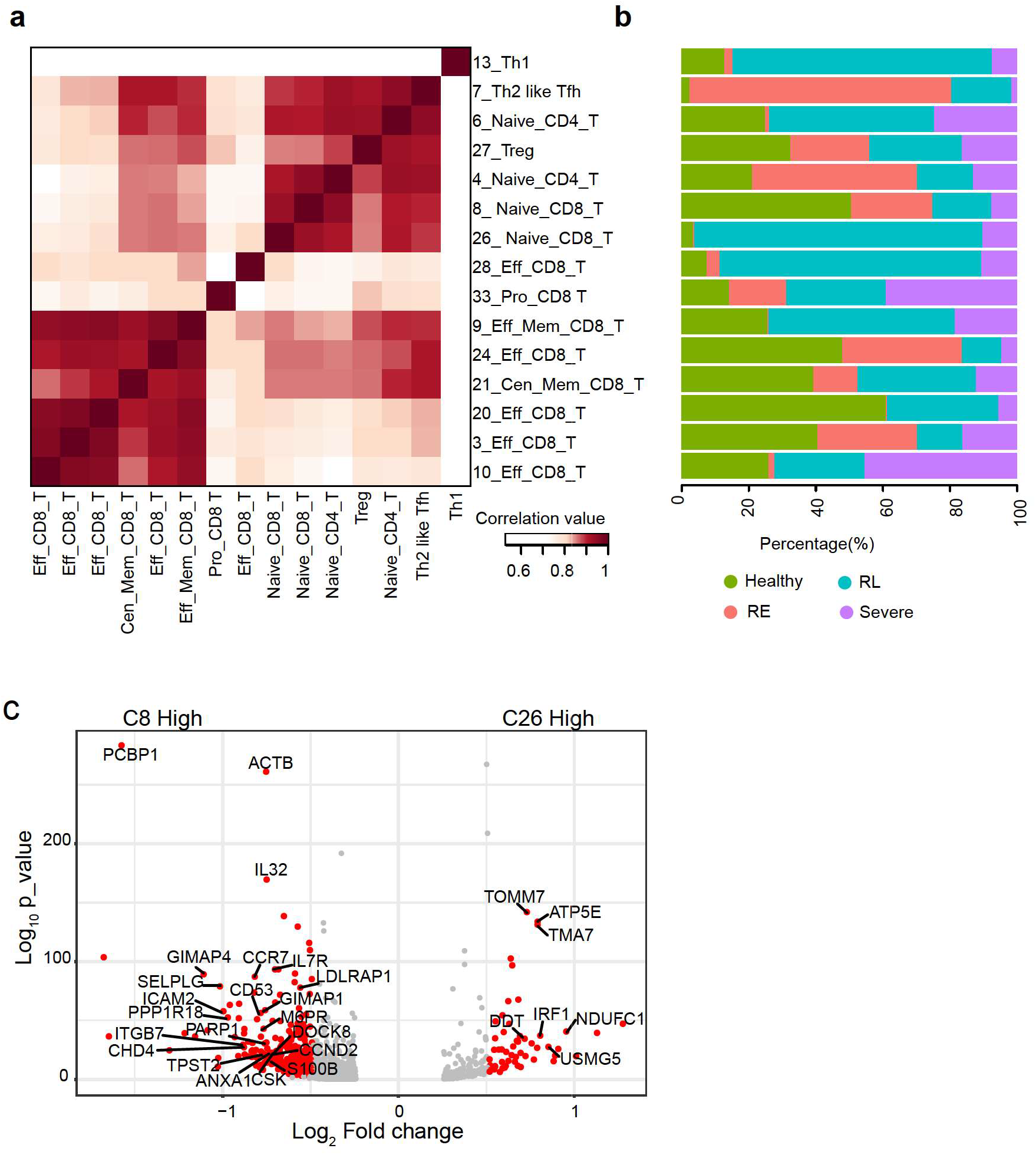
Clustering of single cells using scRNA-seq data. a. Heatmap showing the correlations of the clusters of T lymphocytes. b. Barplot showing the composition of Healthy, RE, RL and Severe patients in the clusters of a. c. Volcano plot showing the DEGs between cluster 8 and cluster 26 naïve CD8 T cells.

**Figure S5.**
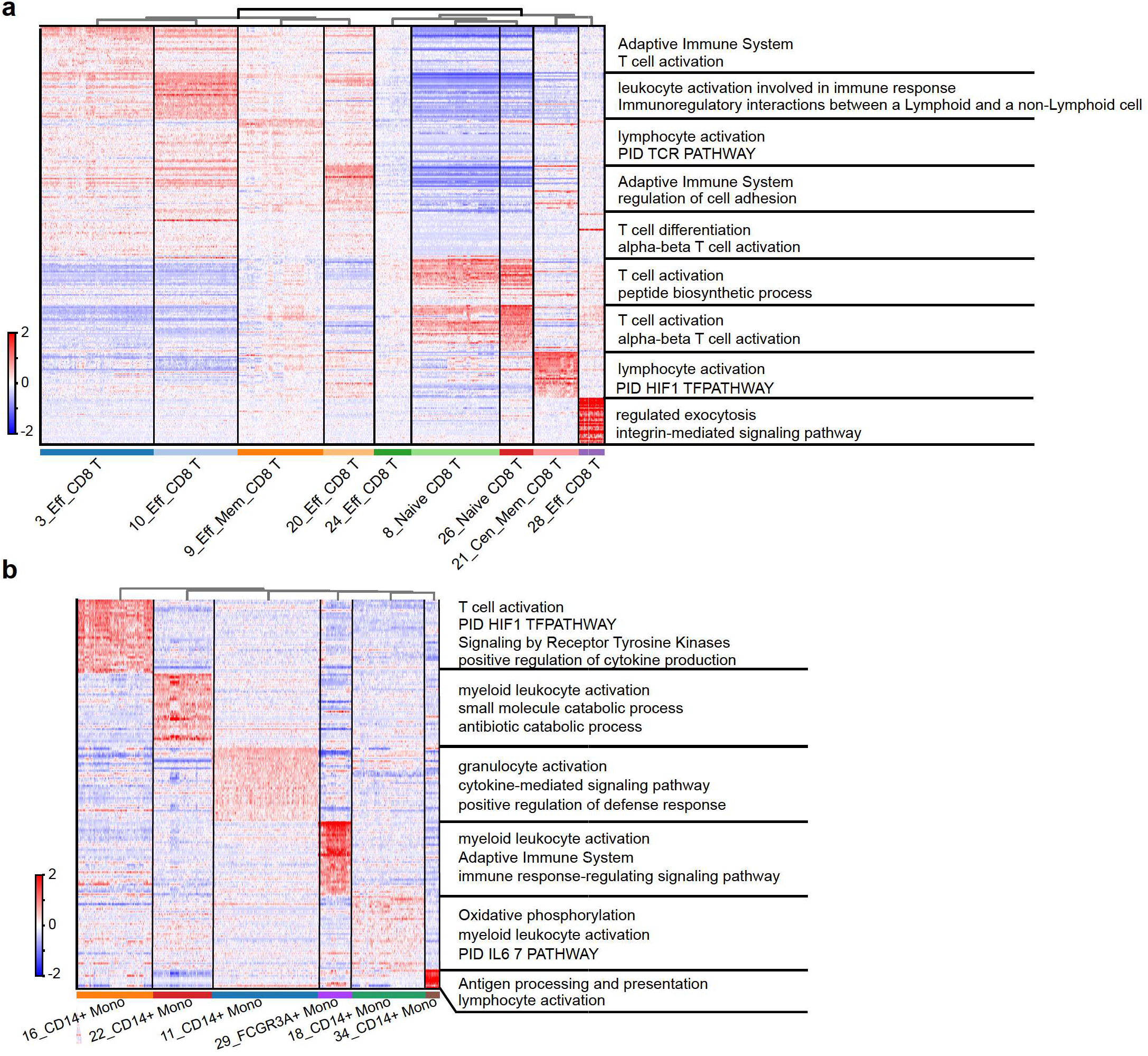
Subtypes of CD8 T cells and monocytes. Heatmap showing the DEGs between the cell subtypes of CD8 T cells (a) and monocytes (b). Representative GO terms are shown on the right.

**Figure S6.**
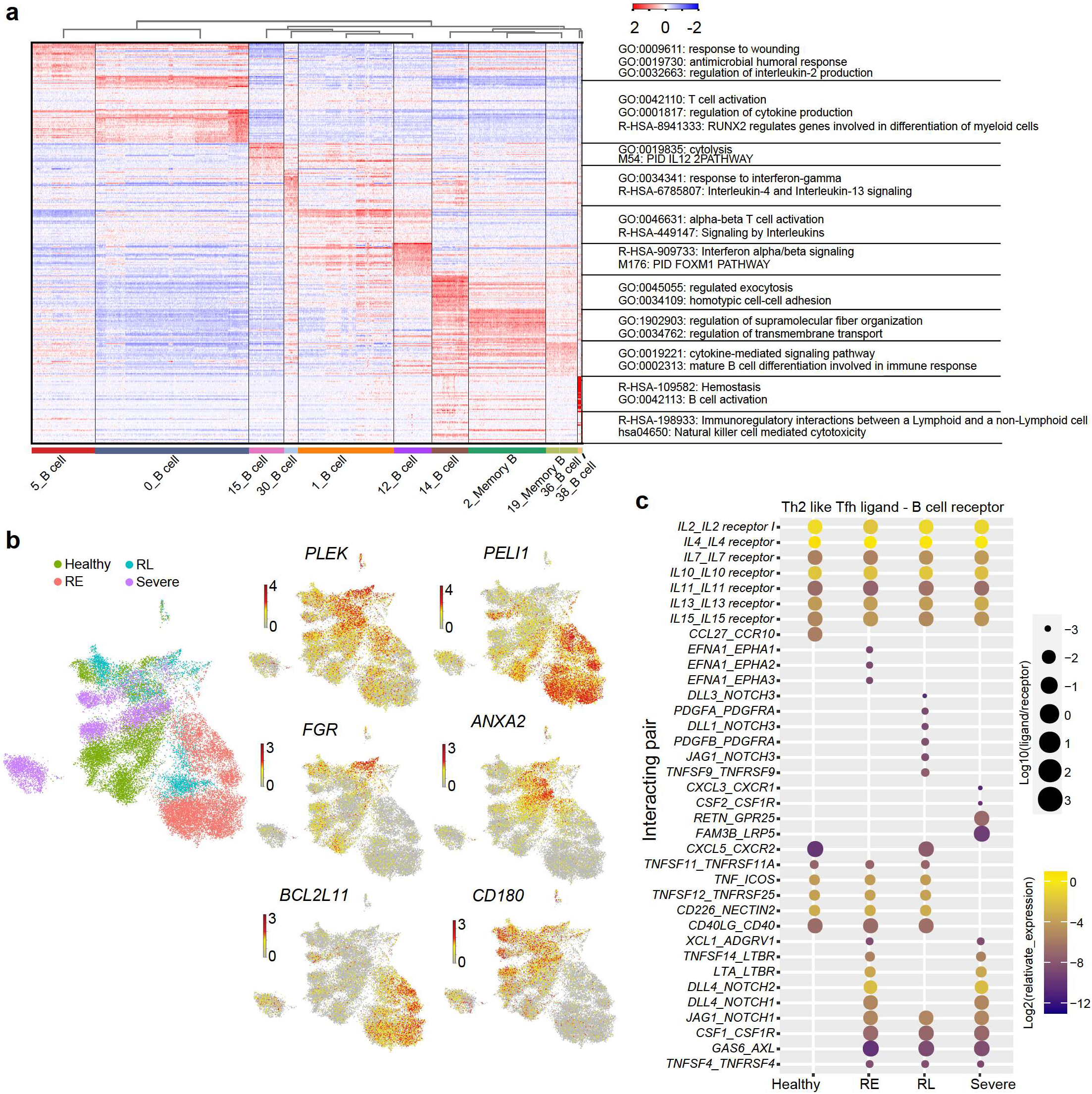
Subtypes of B cells. a. Heatmap showing the DEGs between the cell subtypes of B lymphocytes. Representative GO terms are shown on the right. b. Feature plot showing representative genes of the DEGs in Fig. 2n. c. Interaction of Th2 like Tfh cells (ligand) with B cells in each group of patients.

**Figure S7.**
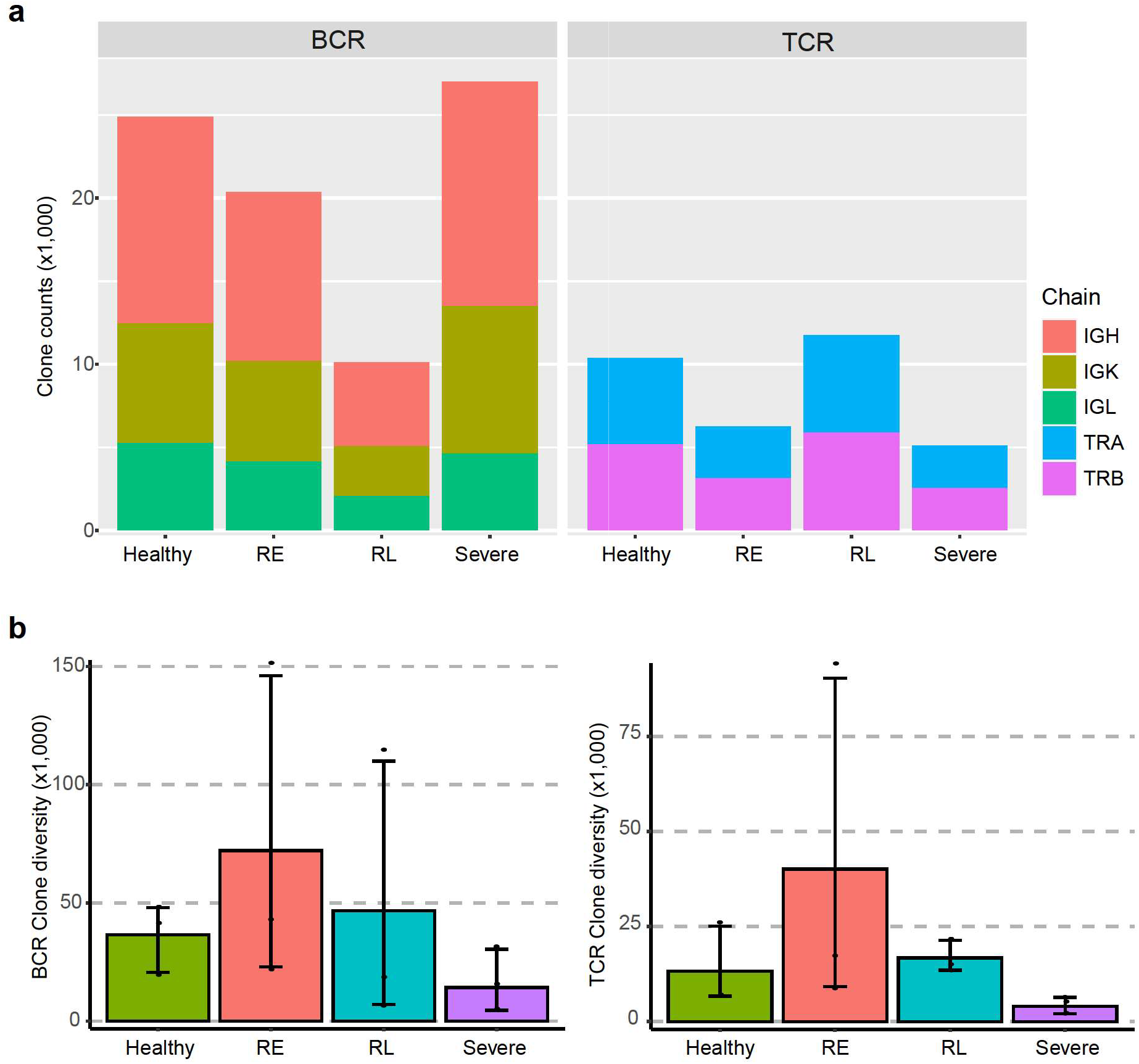
Statistics of BCR and TCR in each group of sample. a. The clone counts observed for each strand of BCR and TCR in Healthy, RE, RL and Severe patients. b. The estimated clone diversity in Healthy, RE, RL and Severe patients.

**Figure S8.**
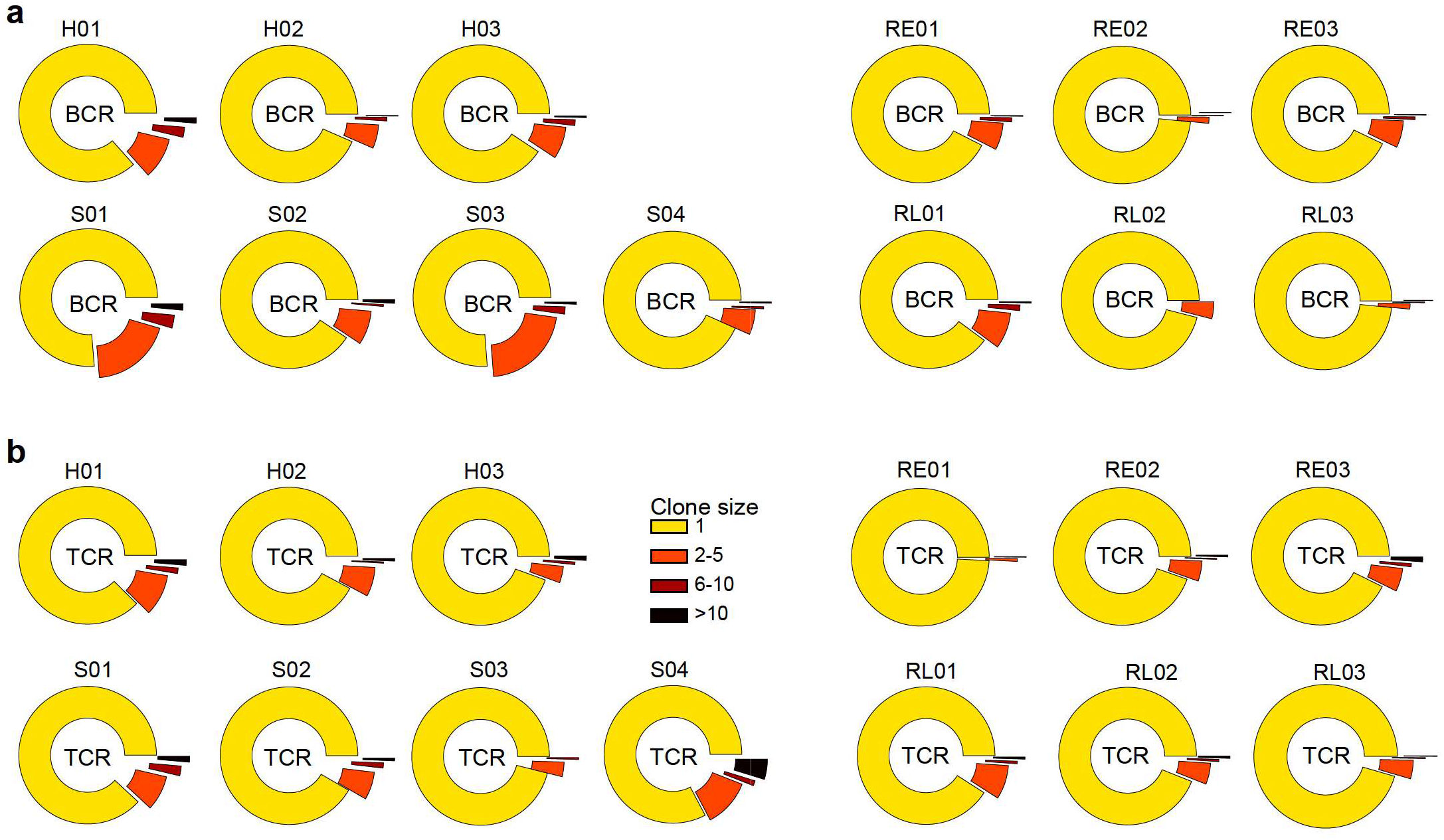
Statistics of BCR and TCR in each group of sample. Calculation of expended clone ratios of BCR and TCR in every individual patient of each group.

**Figure S9.**
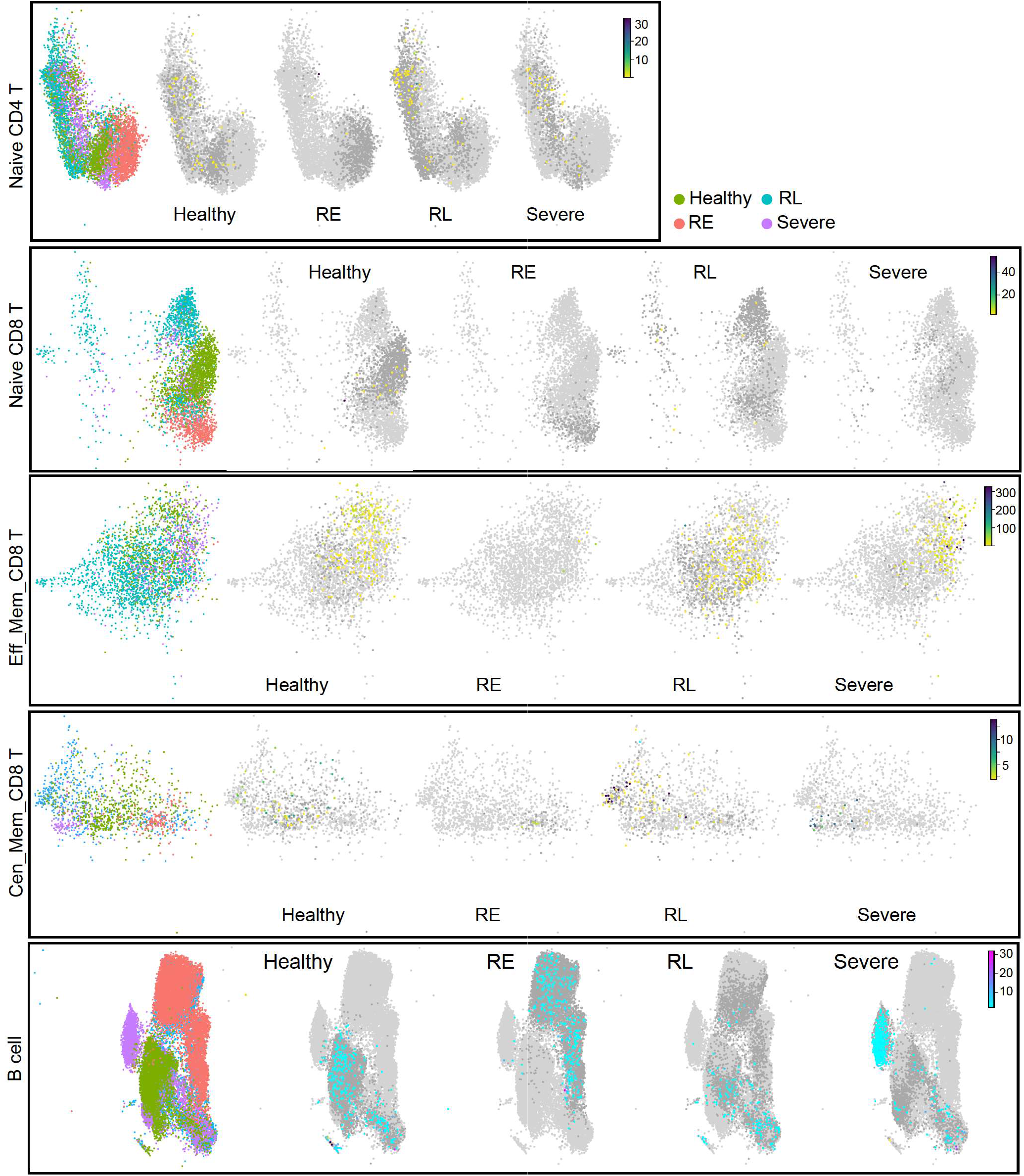
Statistics of BCR and TCR in each group of sample. Feature plot showing the clonal expended cells and their clone size in Naïve CD4 T cells, Naïve CD8 T cells, Effector memory CD 8 T cells, central memory CD 8 T cells and B cells in Healthy, RE, RL and Severe patients. The dark grey dots represent the cells with only one copy of clone detected.

